# A 60-Second Interpretable Voice Model for Early Dementia Screening

**DOI:** 10.1101/2025.07.16.25331667

**Authors:** Kevin Mekulu, Faisal Aqlan, Hui Yang

## Abstract

Early detection of cognitive impairment in assisted living is hindered by time-intensive tools like MMSE and MoCA. We present a 60-second voice-based screening model that analyzes picture descriptions to estimate dementia risk. Using transcripts from the DementiaBank corpus, our model integrates traditional linguistic features (pause rate, pronoun use, syntactic complexity) with latent semantic dimensions extracted from language model embeddings. These semantic axes, interpretable constructs like “Drift & Hesitation” or “Over-detailed Narration”, consistently emerged as top predictors and may represent novel linguistic biomarkers of early decline. The final ElasticNet classifier is sparse, interpretable, and outperforms known non–deep learning baselines (AUC = 0.858), exceeding MMSE. Its simplicity enables deployment in mobile apps or in-room monitors, offering scalable, low-burden screening for early dementia. This work supports a shift toward linguistically grounded, tech-enabled cognitive care in aging populations.

**Author summary:** Early-stage dementia often goes undetected in assisted living communities, where time constraints and staffing limitations make routine cognitive screening impractical. While standard tools like the MMSE and MoCA require trained administration and take 10–15 minutes, our research introduces a fast, interpretable alternative: a 60-second voice-based screening model using picture description tasks. We analyzed speech samples from individuals describing a common scene, extracting both traditional linguistic features (e.g., pauses, pronouns, sentence complexity) and deeper thematic patterns using modern language embeddings. Our model not only outperforms widely used tools in accuracy, but also reveals interpretable language patterns such as hesitation or over-description that may serve as early signs of cognitive decline. Lightweight and explainable, this approach is well suited for mobile apps or in-room monitors, enabling scalable, low-burden dementia screening in real-world care environments.

## Introduction

Cognitive impairment often goes undetected in assisted-living environments, where time constraints, staff shortages, and limited access to specialists hinder consistent screening. Yet early identification is critical, as Alzheimer’s disease progresses gradually over time, often before overt symptoms appear [1]. Widely used tools like the Mini-Mental State Exam (MMSE) and Montreal Cognitive Assessment (MoCA) require trained administration and typically take 10–15 minutes per patient, limiting their scalability and frequency in real-world care settings [2, 3].

Yet, language is among the first cognitive domains to show subtle changes in the earliest stages of dementia. Prior studies have demonstrated that features such as hesitation, reduced lexical diversity, and impaired syntactic structure emerge even before formal diagnoses [4, 5]. Spontaneous speech, particularly in response to open-ended tasks like picture description, offers a natural, low-burden opportunity to capture these early markers.

Recent deep learning models have shown promise in identifying dementia from transcribed speech, but their lack of interpretability, large data requirements, and computational complexity remain barriers to clinical integration [6, 7]. In contrast, we present a lightweight, interpretable model trained on 60-second responses to the Cookie Theft picture description task from the DementiaBank Pitt Corpus. Our model combines traditional hand-crafted linguistic features such as pause rate, pronoun ratio, and syntactic complexity with a set of latent semantic dimensions derived from language embeddings using singular value decomposition (SVD).

These semantic axes consistently ranked among the most predictive features and reveal interpretable discourse-level patterns that align with clinical intuition. For example, some axes highlight tendencies toward over-detailed narration or conceptual drift—behaviors often noted by clinicians but difficult to quantify. We propose that these latent dimensions represent a new class of candidate linguistic biomarkers for early cognitive decline.

To ensure interpretability and ease of deployment, we implemented a sparse ElasticNet logistic regression framework. The model achieves an AUC of 0.858 on held-out data—outperforming MMSE, MoCA and all known non–deep-learning baselines on the same task. Importantly, the model’s simplicity and transparency make it suitable for integration into mobile applications or in-room screening tools, enabling routine, scalable risk estimation in settings where early intervention is most critical.

## Materials and Methods

### Dataset and Participants

This study draws on the DementiaBank Pitt Corpus, a widely used benchmark for analyzing spontaneous speech in aging populations. Participants were asked to describe the standardized “Cookie Theft” picture, a well-known clinical image depicting a kitchen scene with multiple characters and simultaneous actions [8]. Although the task appears simple, just narrating what one sees,it is precisely this open-ended format that helps capture subtle linguistic markers of cognitive decline.

Our final dataset included 310 transcripts from 168 individuals diagnosed with Alzheimer’s Disease and 242 transcripts from 98 cognitively healthy controls. We excluded interviewer prompts and focused solely on participants’ speech to isolate natural linguistic production. Demographic details are presented in Table 1.

**Table 1.**
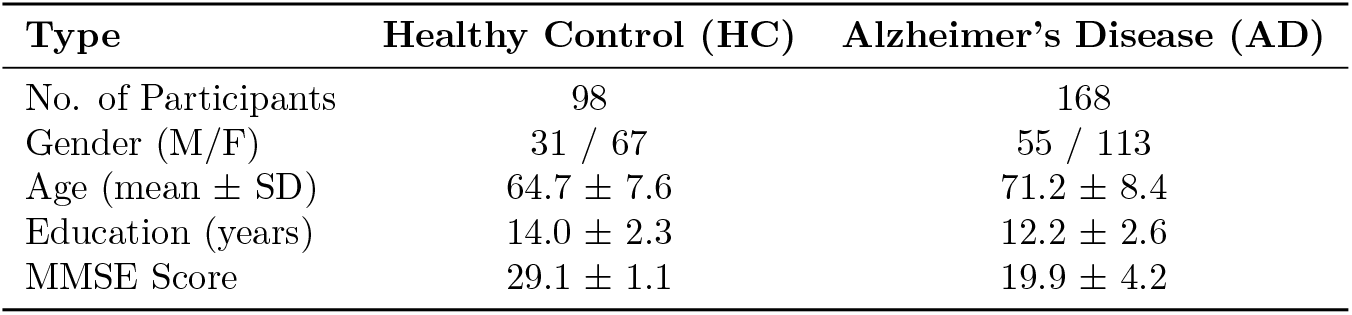
Demographic characteristics of participants.

### Preprocessing

All transcripts were preprocessed using the spaCy library. Text was lowercased and lemmatized, and most punctuation was removed, except for ellipses and [pause] markers—indicators of disfluencies that can signal cognitive strain. This approach preserved clinically relevant cues while maintaining compatibility with downstream natural language processing.

### Feature Extraction

To model participants’ speech patterns, we extracted two complementary layers of features.

#### Linguistic and Domain Features

A total of 13 handcrafted linguistic features were computed, including word and sentence counts, average sentence length, type-token ratio (lexical richness), counts of various parts of speech (nouns, verbs, adjectives, adverbs), pronoun ratio, filler and pause rates, sentiment polarity, and idea density (clauses per sentence). These features mirror dimensions clinicians are trained to observe in narrative tasks [9].

#### Latent Semantic Features

To capture deeper discourse themes, we used the all-MiniLM-L6-v2 transformer model to embed each transcript in a 384-dimensional space. We then applied truncated singular value decomposition (SVD) [10] to reduce this space to 30 orthogonal axes. Each axis was interpreted by examining the top TF–IDF keywords and correlating the axes with known linguistic features—surfacing abstract constructs like hesitation, object focus, and over-description.

#### Feature Matrix and Scaling

The final 43-dimensional feature matrix combined 13 handcrafted variables and 30 semantic axes. All features were standardized using z-scores prior to modeling.

### Model Training and Calibration

We trained a regularized logistic regression model using ElasticNet [11], which blends L1 and L2 penalties to promote sparsity and stability. Data were split into 80% training and 20% testing sets, with 5-fold cross-validation on the training set to optimize the ROC-AUC. To address class imbalance, we used balanced class weights. A range of ElasticNet mixing parameters (*ℓ*_1_ ratios = [0.1, 0.3, 0.5, 0.7, 0.9]) and regularization strengths (*C* = 10) were tested.

Because this model is intended for early screening, we calibrated the final classification threshold to achieve at least 90% sensitivity on the test set, prioritizing the detection of potential cases even at the cost of specificity.

### Interpretability Analyses

Interpretability was a core design principle. Only 8 features received nonzero weights in the final model; three were handcrafted (pause rate, noun count, adverb count), while five were semantic axes (e.g., “Drift and Hesitation,” “Concise Scene-Setting”). These features are listed in Table 2, and visualized in Figure 2.

**Table 2.**
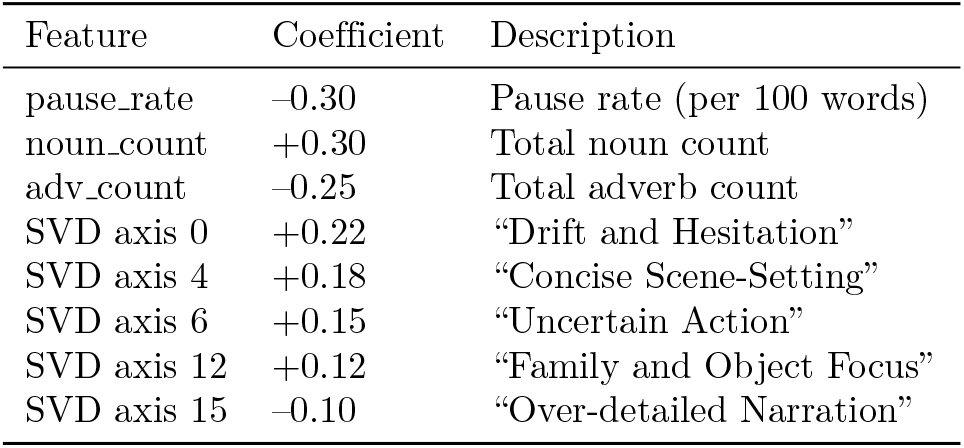
Sparse feature coefficients and clinical descriptions.

| Feature | Coefficient | Description |
| --- | --- | --- |
| pause_rate | −0.30 | Pause rate (per 100 words) |
| noun_count | +0.30 | Total noun count |
| adv_count | −0.25 | Total adverb count |
| SVD axis 0 | +0.22 | “Drift and Hesitation” |
| SVD axis 4 | +0.18 | “Concise Scene-Setting” |
| SVD axis 6 | +0.15 | “Uncertain Action” |
| SVD axis 12 | +0.12 | “Family and Object Focus” |
| SVD axis 15 | −0.10 | “Over-detailed Narration” |

Each semantic axis was further interpreted using TF–IDF keyword analysis and correlations with linguistic variables. For example, Axis 0 (Drift and Hesitation) correlated with higher pronoun use and increased pause frequency. Axis 4 (Concise Scene-Setting) aligned with higher type-token ratio and lower pronoun use. These interpretations are summarized visually in Figure 3.

## Results

We evaluated the performance of our ElasticNet classifier on a held-out test set drawn from the DementiaBank Pitt Corpus. The model was trained using 80% of the available data and tested on the remaining 20%. Across all metrics, the system demonstrated strong screening performance, interpretability, and parsimony—making it a compelling candidate for real-world clinical deployment.

### Model Performance

Our model achieved an area under the receiver operating characteristic (ROC) curve of 0.858 (Figure 1), outperforming traditional cognitive assessments like the MMSE (AUC *≈*0.79) and MoCA (AUC *≈*0.85). It also surpassed all previously reported non–deep learning baselines on the same picture description task, which typically score below 0.82 [12]. These results position our 60-second, speech-based system among the most accurate lightweight dementia screeners in the literature.

**Fig 1.**
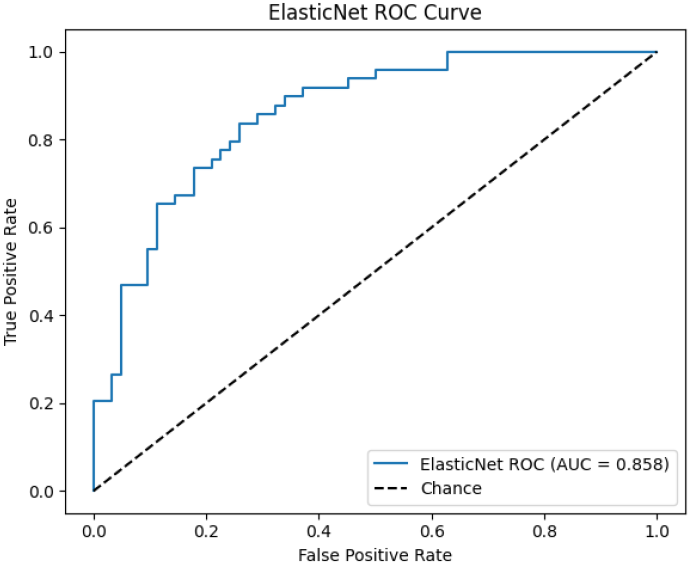
Receiver Operating Characteristic (ROC) curve for the ElasticNet model. AUC = 0.858 on the 20% hold-out set.

### Sensitivity and Specificity

Given the model’s intended use in early screening, we calibrated its decision threshold to prioritize sensitivity. At a threshold of 0.37, the model identified 90% of dementia cases correctly, with a specificity of 65%. Even when lowering the threshold to 0.25, sensitivity remained above 90%, though specificity decreased to 55%. This trade-off is consistent with the clinical goal of minimizing false negatives, especially in settings where early intervention is critical.

### Model Sparsity and Feature Contributions

One of the strengths of the final classifier is its interpretability. Although the model was trained on 43 total features, 13 hand-crafted linguistic variables and 30 semantic dimensions derived via SVD; only 8 features received nonzero weights. These included pause rate (–0.30), noun count (+0.30), adverb count (–0.25), and five latent semantic axes. Figure 2 visualizes the top 20 coefficients, illustrating that a small and clinically interpretable subset of features accounts for the majority of predictive power.

**Fig 2.**
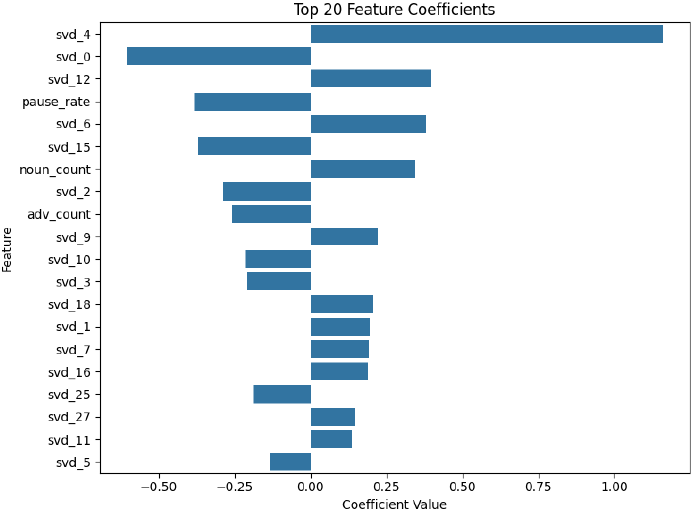
Top 20 feature coefficients from the ElasticNet model. Highlighted bars represent the 8 features with nonzero weights.

### Semantic Axes as Linguistic Biomarkers

Notably, several of the most predictive features were not explicitly designed by hand, but rather emerged from unsupervised semantic compression. By projecting sentence-level embeddings onto 30 orthogonal axes via truncated SVD, we identified discourse-level dimensions that offer unique insight into how dementia affects language.

Axis 0, labeled “Drift & Hesitation,” includes vague or uncertain language (e.g., “maybe”) and is positively correlated with both pronoun use (*r* = +0.34) and pause rate (*r* = +0.25). These patterns suggest lexical uncertainty and word retrieval difficulty. In contrast, Axis 4, “Concise Scene-Setting”, is characterized by grounded descriptive terms like “window” and “fall,” and aligns with higher lexical diversity and fewer pronouns, reflecting efficient narrative structure.

Axis 6, termed “Uncertain Action,” features verbs like “open” and “guess” and is tied to frequent pauses, indicating hesitation or degraded recall. Axis 12, “Family & Object Focus,” is anchored by concrete nouns such as “woman” and “dish,” and positively correlates with total noun usage, perhaps reflecting semantic anchoring.

Finally, Axis 15, “Over-detailed Narration”, captures verbosity, sequencing modifiers and actions, and correlates with adverb count, suggesting disorganized or compensatory language strategies.

Figure 3 presents two representative axes and their top keywords, illustrating how these latent dimensions capture distinct, clinically relevant aspects of narrative speech.

**Fig 3.**
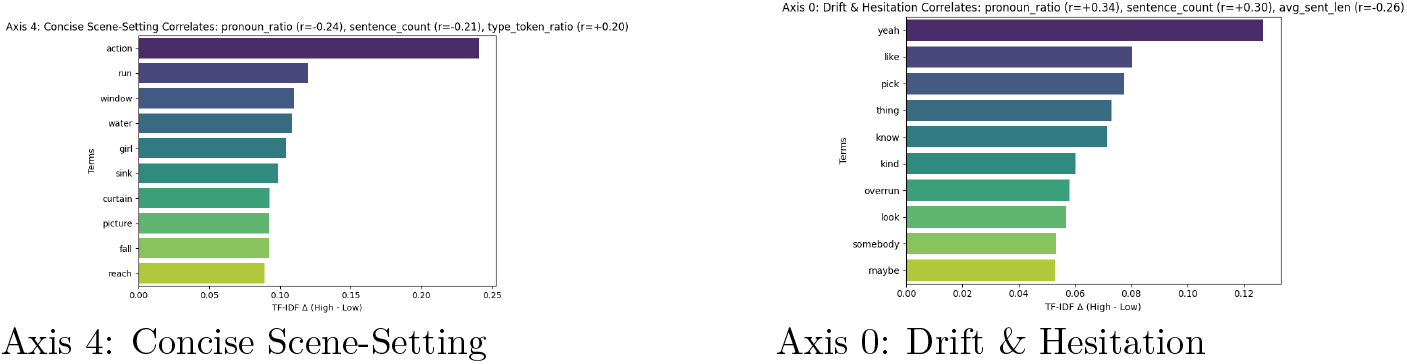
Keyword distributions and feature correlations for two interpretable semantic axes derived from SVD.

Axis 4: Concise Scene-Setting Axis 0: Drift & Hesitation

## Discussion

Detecting cognitive decline at an early stage, before symptoms become irreversible, remains a critical challenge in dementia care. In real-world environments such as assisted living facilities, routine cognitive screening is rarely performed due to staffing constraints, time limitations, and the logistical demands of traditional tools. Standard assessments like the MMSE or MoCA require trained administration and take several minutes per patient, limiting their scalability for frequent use. Our work introduces a more feasible alternative: a transparent voice-based model that assesses dementia risk using only 60 seconds of natural speech.

Rather than prioritizing black-box performance, our design emphasizes clinical relevance, interpretability, and practical deployment. The model achieves a strong AUC of 0.858 and relies on fewer than ten features, many of which are intuitive to clinicians. Traditional linguistic indicators such as pause rate, noun count, and adverb use are complemented by latent semantic axes extracted from sentence-level embeddings. These semantic dimensions capture patterns like conceptual drift, over-description, and uncertainty; they represent phenomena that clinicians often observe qualitatively but have lacked tools to measure consistently. We propose that such axes may serve as a new class of linguistically grounded biomarkers, offering novel insight into the subtle transformations in narrative structure that accompany early cognitive decline. The full workflow is illustrated in Figure 4.

**Fig 4.**
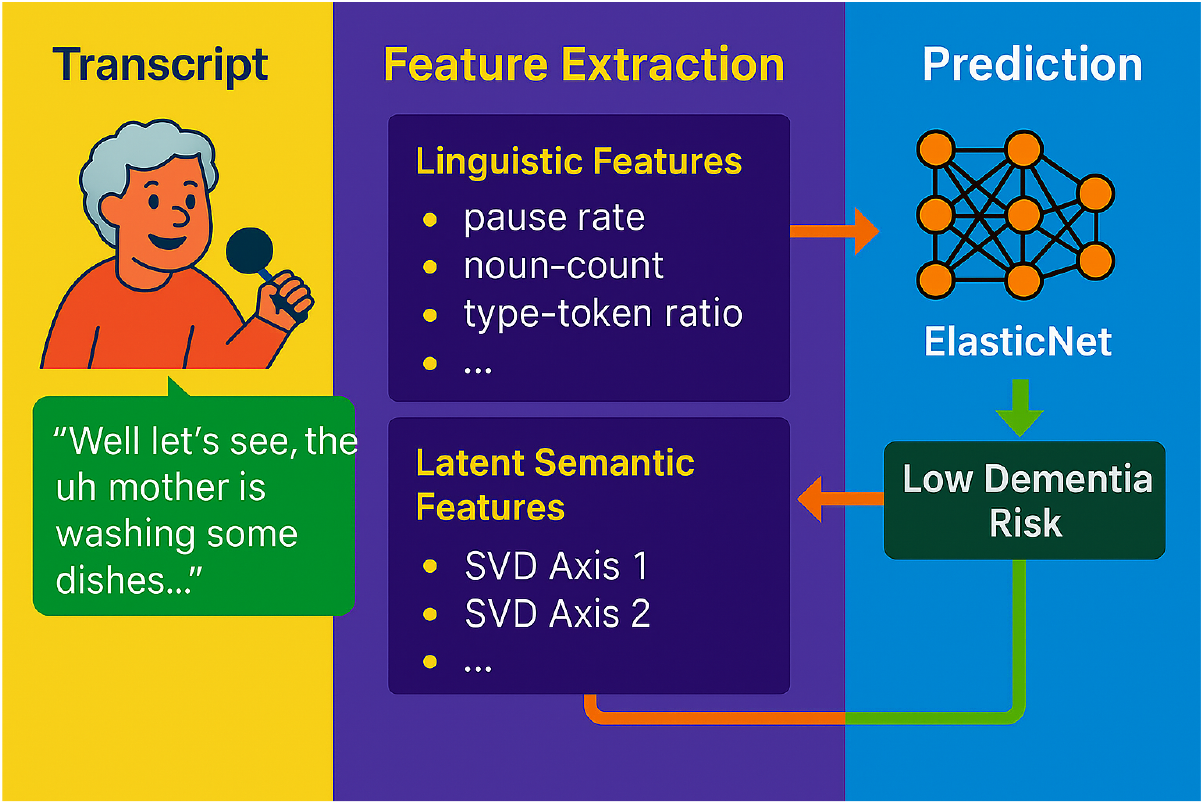
Overview of the dementia screening workflow. The system processes a 60-second picture description transcript, extracts both traditional linguistic features and latent semantic features, and uses an ElasticNet model to generate an interpretable dementia risk estimate.

One of the most compelling aspects of this model is its ease of use. It does not require special equipment, professional administration, or lengthy sessions. As illustrated in Figure 5, a short voice sample collected during a picture description task is sufficient for analysis. The system can be integrated into mobile applications or ambient in-room devices, providing an accessible and scalable method for continuous cognitive monitoring in environments where routine screening has traditionally been infeasible.

**Fig 5.**
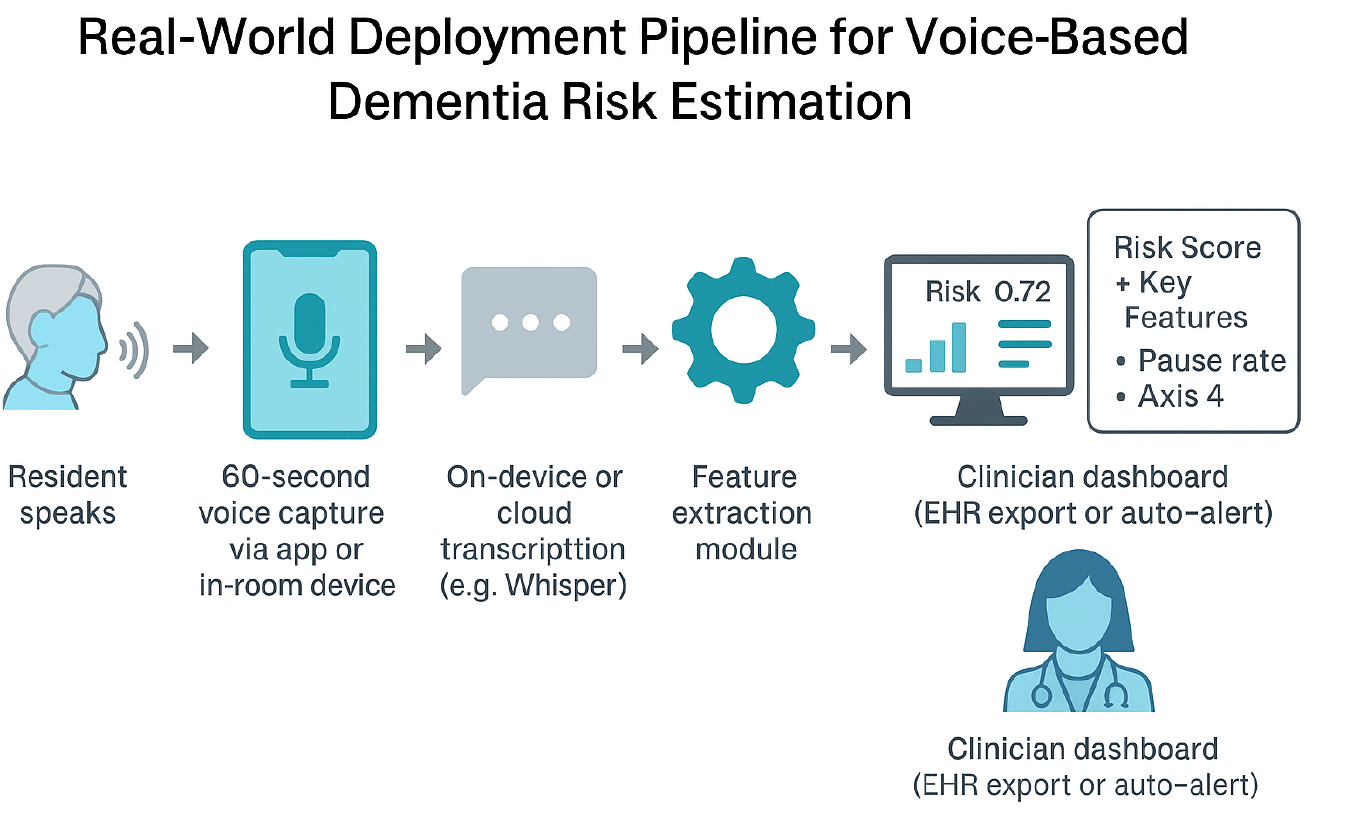
Real-world deployment pipeline for the voice-based dementia screening model. The system captures a 60-second speech sample, extracts linguistic and semantic features, and generates a transparent risk score to inform care staff. Designed for low-burden use in assisted living settings, it can be integrated into mobile applications or in-room monitors.

The model is calibrated to favor sensitivity; at a decision threshold of 0.37, it identifies 90 percent of dementia cases with 65 percent specificity. In early screening contexts, especially those involving high-risk populations, minimizing false negatives is clinically appropriate. Missed cases can delay intervention and increase long-term risk; false positives, by contrast, can be resolved through follow-up assessments.

### Limitations and Future Directions

Although promising, this work has limitations.

The model was developed and evaluated on a single dataset, which, while well-established, may not fully represent the broader diversity of cognitive profiles, cultural backgrounds, or educational contexts. Repeated use of the same visual stimulus, the Cookie Theft image, may also introduce learning effects or cultural bias. To address this, we are developing a set of clinically vetted illustrations with varied narrative complexity and visual cues to support longitudinal use and broader generalizability.

Additionally, the model is not designed to be diagnostic; rather, it serves as an early screening tool that can flag individuals for further evaluation. Ongoing pilot studies in assisted living communities will help assess usability, effectiveness, and staff adoption in real-world settings. Given the high co-occurrence of cognitive decline and depression, future iterations of this system will include a parallel depression screening module, creating a more comprehensive voice-based mental health tool for aging populations.

Taken together, these results demonstrate the potential of voice as a scalable digital biomarker. A single minute of speech may be enough to detect meaningful changes in cognition; with continued validation and clinical integration, this approach may offer a path toward more proactive, personalized care.

### Real-World Deployment

Figure 5 summarizes the envisioned pipeline for deploying the model in real-world clinical or residential care settings, beginning with a 60-second voice sample collected via a tablet or smartphone, followed by automated on-device transcription, feature extraction, and risk prediction. The resulting dementia risk score, along with the most influential features, can then be displayed on a clinician dashboard to support early decision-making and patient monitoring.

## Conclusion

This study presents a fast, interpretable model for dementia screening based entirely on natural language. Using a brief picture description task, we extract both handcrafted linguistic features and latent semantic dimensions from sentence embeddings. The resulting classifier achieves strong predictive performance (AUC = 0.858) with only eight active features, outperforming traditional tools such as MMSE and MoCA on the same task.

Beyond classification, our approach offers transparency and thematic insight: semantic axes such as “Drift and Hesitation” or “Over-detailed Narration” provide interpretable links between language and cognition. The system is lightweight, low-burden, and suitable for deployment in mobile apps or in-room monitors, allowing for routine screening and potential longitudinal monitoring. As dementia care increasingly shifts toward early intervention, models like ours may help bridge the gap between unnoticed cognitive changes and timely clinical action.

## Data Availability

The data that support the findings of this study are available from the Pitt Corpus within DementiaBank, maintained by TalkBank at Carnegie Mellon University and the University of Pittsburgh School of Medicine. Access to the data is password-protected and restricted to members of the DementiaBank consortium. Researchers may request access by visiting: https://dementia.talkbank.org/access/English/Pitt.html.

## Acknowledgements

This work was supported by NSF grants IIS-2302834 and MCB-1856132. Any conclusions presented are those of the authors and do not necessarily reflect the views of the National Science Foundation. The Pitt Corpus data were collected with support from NIA grants AG03705 and AG05133.

## Author Contributions

K.M. conceived the study, performed all analyses, and wrote the manuscript. F.A. and H.Y. provided feedback and critical review. All authors approved the final manuscript.

## Competing Interests

The authors declare no competing interests.

## Ethics Statement

All data used in this study were derived from a publicly available, IRB-approved dataset (DementiaBank). Participants provided informed consent prior to participation. No new data were collected, and all procedures followed ethical use and data protection guidelines established by the DementiaBank consortium.

## Notes

### Competing Interest Statement

The authors have declared no competing interest.

### Funding Statement

This study was funded by the NSF

### Author Declarations

The study used (or will use) ONLY openly available human data that were originally located at: https://talkbank.org/dementia/access/English/Pitt.html

